# Genetic associations and architecture of asthma-chronic obstructive pulmonary disease overlap

**DOI:** 10.1101/2020.11.26.20236760

**Authors:** C. John, A.L. Guyatt, N. Shrine, R. Packer, T.A. Olafsdottir, J. Liu, L.P. Hayden, S.H. Chu, J.T. Koskela, J. Luan, X. Li, N. Terzikhan, H. Xu, T.M. Bartz, H. Petersen, S. Leng, S.A. Belinsky, A. Cepelis, A.I. Hernández Cordero, M. Obeidat, G. Thorleifsson, D.A. Meyers, E.R. Bleecker, L.C. Sakoda, C. Iribarren, Y. Tesfaigzi, S.A. Gharib, J. Dupuis, G. Brusselle, L. Lahousse, V.E. Ortega, I. Jonsdottir, D. D. Sin, Y. Bossé, M. van den Berge, D. Nickle, J.K. Quint, I. Sayers, I.P. Hall, C. Langenberg, S. Ripatti, T. Laitinen, A.C. Wu, J. Lasky-Su, P. Bakke, A. Gulsvik, C.P. Hersh, C. Hayward, A. Langhammer, B. Brumpton, K. Stefansson, M.H. Cho, L.V. Wain, M.D. Tobin

**Affiliations:** Department of Health Sciences, University of Leicester, Leicester, UK; deCODE genetics/Amgen, Reykjavik, Iceland; Channing Division of Network Medicine, Brigham and Women’s Hospital and Harvard Medical School, Boston, USA; Institute for Molecular Medicine Finland, University of Helsinki, Helsinki, Finland; MRC Epidemiology Unit, University of Cambridge School of Clinical Medicine, Cambridge, UK; Division of Genetics, Genomics and Precision Medicine, Department of Medicine, University of Arizona, Tucson, USA; Department of Epidemiology, Erasmus Medical Center, Rotterdam, Netherlands; Department of Biostatistics, Boston University School of Public Health, Boston, USA; Cardiovascular Health Research Unit, Department of Medicine & Department of Biostatistics, University of Washington, Seattle, USA; Lovelace Respiratory Research Institute, Albuquerque, USA; Division of Epidemiology, Biostatistics, and Preventive Medicine, Department of Internal Medicine, University of New Mexico, Albuquerque, USA; Department of Public Health and Nursing, Faculty of Medicine and Health Sciences, Norwegian University of Science and Technology (NTNU), Levanger, Norway; Centre for Heart Lung Innovation, University of British Columbia, Vancouver, British Columbia, Canada; Faculty of Medicine, School of Health Sciences, University of Iceland, Reykjavik, Iceland; Division of Research, Kaiser Permanente of Northern California, Oakland, USA; Brigham and Women’s Hospital and Harvard Medical School, Boston, USA; Computational Medicine Core, Center for Lung Biology & UW Medicine Sleep Center, Medicine, University of Washington, Seattle, USA; Department of Respiratory Medicine, Ghent University Hospital, Ghent, Belgium; Department of Bioanalysis, Ghent University, Ghent, Belgium; Department of Medicine, Wake Forest School of Medicine, Winston-Salem, USA; Institut universitaire de cardiologie et de pneumologie de Québec, Department of Molecular Medicine, Laval University, Québec, Canada; University of Groningen, University Medical Center Groningen, Department of Pulmonology, GRIAC Research Institute, Groningen, The Netherlands; Global Health, University of Washington, Seattle, USA; Gossamer Bio, San Diego, USA; National Heart and Lung Institute, Imperial College London, London, UK; Division of Respiratory Medicine and NIHR Nottingham Biomedical Research Centre, University of Nottingham, Nottingham, UK; Biodiscovery Institute, University of Nottingham, Nottingham, UK; Broad Institute of MIT and Harvard, Cambridge, USA; Division of Medicine, Department of Pulmonary Diseases, Turku University Hospital, Turku, Finland; Department of Pulmonary Diseases and Clinical Allergology, University of Turku, Turku, Finland; Center for Healthcare Research in Pediatrics (CHeRP) and PRecisiOn Medicine Translational Research (PROMoTeR) Center, Department of Population Medicine, Harvard Pilgrim Health Care Institute and Harvard Medical School, Boston, USA; Department of Clinical Science, University of Bergen, Bergen, Norway; MRC Human Genetics Unit, Institute of Genetics and Molecular Medicine, University of Edinburgh, Edinburgh, UK; K.G. Jebsen Center for Genetic Epidemiology, Department of Public Health and Nursing, Norwegian University of Science and Technology (NTNU), Trondheim, Norway; Clinic of Thoracic and Occupational Medicine, St. Olav’s Hospital, Trondheim University Hospital, Trondheim, Norway; Leicester NIHR Biomedical Research Centre, Leicester, UK

## Abstract

Some individuals have characteristics of both asthma and COPD (asthma-COPD overlap, ACO), and evidence suggests they experience worse outcomes than those with either condition alone. Improved knowledge of the genetic architecture would contribute to understanding whether determinants of risk in this group differ from those in COPD or asthma.

We conducted a genome-wide association study in 8,068 cases and 40,360 controls of European ancestry from UK Biobank (stage 1). After excluding variants only associated with asthma or COPD we selected 31 variants for further investigation in 12 additional cohorts (stage 2), and discovered eight novel signals for ACO in a meta-analysis of stage 1 and 2 studies.

Our signals include an intergenic signal on chromosome 5 not previously associated with asthma, COPD or lung function, and suggest a spectrum of shared and distinct genetic influences in asthma, COPD and ACO. A number of signals may represent loci that predispose to serious long-term consequences in people with asthma.

## Introduction

Chronic respiratory diseases are a common and important cause of morbidity and mortality worldwide, affecting over 500 million people and causing 7% of deaths globally.^1^ Chronic obstructive pulmonary disease (COPD) accounts for 55% of chronic respiratory disease prevalence and over 80% of deaths, whilst asthma is the next most common condition, and is particularly prevalent amongst children.^1^ Prevalence is highest in high-income countries (including the USA and Western Europe), though deaths are higher elsewhere, notably in Asia and Oceania.^1^ Globally, chronic respiratory disease is the third leading cause of death.^1^

Asthma and COPD are heterogeneous conditions^2-4^ that share some common features, including airflow obstruction with differing degrees of reversibility. Inflammatory processes are important in the pathogenesis of both conditions, and cytokine profiles, for example, identify both distinct and overlapping groups of patients.^5^ Individuals with characteristics of both conditions have until recently been referred to as having “asthma-COPD overlap” (ACO),^4^ and a number of studies have suggested that patients who meet criteria for both asthma and COPD have significantly worse outcomes than those with either condition alone.^6-13^ More recent guidelines emphasize that asthma and COPD are different conditions, but recognise that they may coexist in the same patient.^14^ Such individuals with features of both diseases risk being excluded from research studies that might provide evidence about the most effective treatment strategies for this group.^3^

Environmental risk factors – notably tobacco smoke in the case of COPD – are central, but genetics also plays an important role in both asthma and COPD.^15-17^ Genome-wide association studies (GWAS) examine variants across the genome agnostically, with the purpose of identifying variant-trait associations that inform understanding of disease biology, and by extension, potential treatment strategies. GWAS have identified a considerable number of loci associated with risk of asthma or COPD in European populations.^18-61^ The genetic correlation (r_g_) between asthma and COPD is 0.38 (p=6.2×10^−5^), suggesting a shared genetic aetiology.^51^ Some GWAS have specifically studied ACO, including a GWAS of ACO compared to COPD alone,^8^ and a meta-analysis of an asthma and COPD GWAS.^62^ At least twenty loci outside of the HLA (human leucocyte antigen) region have been identified as associated with both asthma and lung function or COPD at a threshold of p<5×10^−8^, but have not been specifically described as ACO loci: these include signals in/near *IL1RL1* on chromosome 2,^33, 63^ *STAT6* on chromosome 12,^34, 41^ and *GSDMB/THRA* on chromosome 17.^38, 56^ There are also a number of overlapping loci in the HLA region, including the first shared signal to be identified, *HLA-DQB1/HLA-DQA2*.^30^ A previous GWAS did not identify any variants associated with ACO at the conventional p<5×10^−8^ threshold, but this study was of a modest sample size (n=3570) and thus likely underpowered.^8^

Improved knowledge of genetic variants associated with co-existing asthma and COPD would contribute to understanding of the underlying molecular pathways, clarifying whether the genetic determinants of illness in this patient group are distinct from those in COPD or asthma alone. This knowledge could also help to determine specific management strategies for those with co-existing asthma and COPD. Notwithstanding the controversies of changing terminology for individuals affected with both asthma and COPD, for brevity in this paper, we refer to this case status as “ACO”. Understanding the extent of the shared genetic background between asthma and COPD may inform future discussion about the terminology used to describe people meeting criteria for both asthma and COPD diagnosis.

Accordingly, using spirometry, self-report and electronic healthcare record (EHR) data to define cases with both asthma and COPD (ACO) and suitable controls, we undertook the largest GWAS of coexisting asthma and COPD to date, including up to 12,369 cases and 88,969 controls, in a two-stage design incorporating 13 studies

## Results

For stage 1 analyses, 8068 individuals were selected from UK Biobank as ACO cases (defined by self-reported asthma and spirometry demonstrating obstruction) and 40,360 as healthy controls free of either asthma or COPD. We undertook two further analyses to determine whether signals were driven by COPD or asthma alone, for prioritisation of signals. For these, another 16,762 individuals were selected as controls with COPD alone (without asthma), and 26,815 as controls with asthma alone (without COPD). The age, sex, smoking status and lung function of the cases and controls for both the primary and sensitivity analyses are shown in **Table 1**. ACO cases were slightly older, on average, than healthy controls, and included a higher proportion of males and ever-smokers. Age, sex and smoking status were included as covariates in the association analysis, in addition to principal components for ancestry.

**Table 1.** Descriptive characteristics of cases and controls included in stage 1 (UK Biobank primary and signal prioritisation analyses)

|  | **ACO cases (n=8068)** | **Healthy controls (n=40360)** | **Controls with COPD but no asthma (n=16762)** | **Controls with asthma but no COPD (n=26815)** |
| --- | --- | --- | --- | --- |
| **Age at recruitment** (median, IQR) | 60 (53-65) | 57 (49-63) | 62 (56-65) | 55 (48-61) |
| **Sex** |  |  |  |  |
| Male | 4179 (51.8%) | 17598 (43.6%) | 9147 (54.6%) | 9703 (36.2%) |
| Female | 3889 (48.2%) | 22762 (56.4%) | 7615 (45.4%) | 17112 (63.8%) |
| **Smoking status** |  |  |  |  |
| Ever smoked | 4367 (54.1%) | 17316 (42.9%) | 11752 (70.1%) | 11231 (41.9%) |
| Never smoked | 3701 (45.9%) | 23044 (57.1%) | 5010 (29.9%) | 15584 (58.1%) |
| **Lung function** (median, IQR) |  |  |  |  |
| FEV<sub>1</sub>/FVC | 0.63 (0.58-0.67) | 0.78 (0.75-0.81) | 0.65 (0.61-0.68) | 0.77 (0.74-0.80) |
| % predicted FEV<sub>1</sub> | 66.1% (56.5%-73.3%) | 97.3% (89.9%-105.6%) | 68.7% (60.0%-74.8%) | 90.8% (81.6%-100.0%) |

After filtering based on minor allele frequency (MAF) greater than 0.01 and imputation quality (INFO) greater than 0.5, a total of 7,693,381 variants were retained for downstream analyses. The LDSC (linkage disequilibrium score) regression intercept was 1.018, indicating that our results were not strongly affected by inflation due to population structure.^64^ This value was used to compute corrected standard errors of the discovery results (**Supplementary Figure 1**).

### ACO shares genetic architecture with other traits

Using LD score regression, we computed genetic correlations between ACO, asthma, moderate-severe asthma, COPD, FEV_1_/FVC and blood eosinophil counts, using results from the current study and from published GWAS in UK Biobank and other studies (**Figure 1, Supplementary Table 4**). We observed genetic correlations (r_g_) of broadly similar magnitude between ACO and COPD (r_g_=0.828, p=3.19×10^−299^), ACO and asthma (r_g_ =0.743, p=6.18×10^−44^), and ACO and FEV_1_/FVC (r_g_=-0.692, p=7.48×10^−33^). The genetic correlation (r_g_) between asthma and FEV_1_/FVC was −0.333 (p=8.71×10^−7^), (i.e. increased risk of asthma was correlated with a lower FEV_1_/FVC). Blood eosinophil count showed a moderate genetic correlation with ACO (r_g_=0.292, p=4.87×10^−11^) that was similar in magnitude to the correlation of eosinophils with asthma (r_g_=0.371, p=3.15×10^−7^), whereas the genetic correlations of eosinophils with FEV_1_/FVC (r_g_ =-0.070, p=0.002) and COPD (r_g_=0.130, p=4.83×10^−6^) were smaller. We additionally computed genetic correlations between ACO and 16 traits related to autoimmune disease, as well as the genetic correlation between ACO and smoking behaviour (**Supplementary Table 4**). After asthma, the next strongest genetic correlation was with eczema (r_g_=0.255, p=0.004), followed by multiple sclerosis (r_g_=0.323, p=0.011).

**Figure 1.**
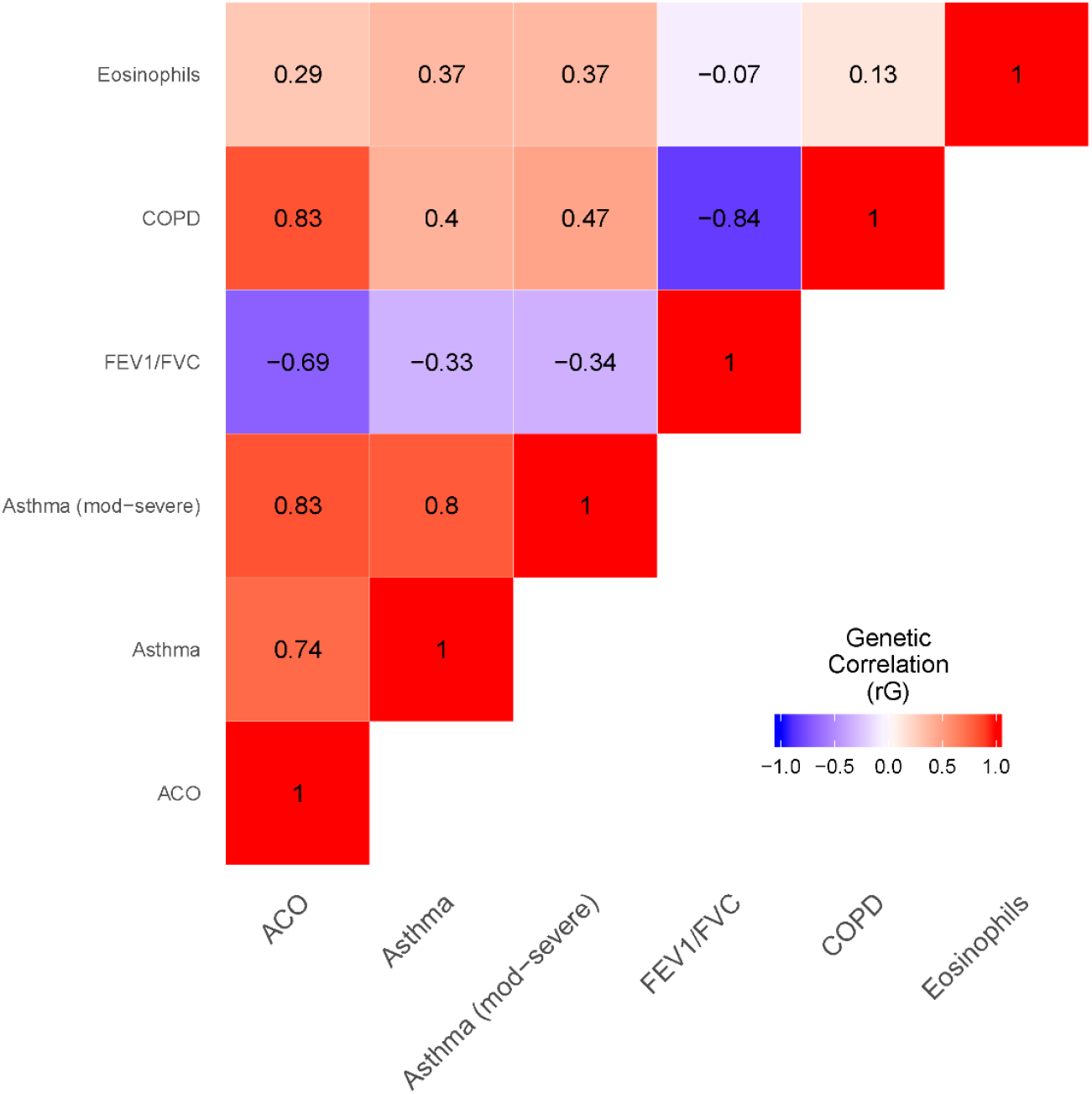
Genetic correlations between ACO and asthma, moderate-severe asthma, COPD, FEV_1_/FVC, and blood eosinophil counts. Genetic correlations were computed using LD score regression. The annotation in each tile represents the magnitude of the genetic correlation estimate (rG), and intensity is proportional to the magnitude of effect. Note that for FEV_1_/FVC, a negative correlation shows that the other trait is associated with reduced FEV_1_/FVC (reduced FEV_1_/FVC implies worse lung function). Datasets used: ACO=current discovery results from UK Biobank; Asthma=GWAS results from Demenais et al., 2018;^34^ Asthma (moderate-severe)=GWAS of asthma by Shrine et al, 2019;^55^ COPD=GWAS of COPD by Sakornsakolpat et al, 2019;^56^ FEV_1_/FVC=GWAS of FEV_1_/FVC (UK Biobank and SpiroMeta) by Shrine et al, 2019;^63^ Eosinophils=blood eosinophil counts published by Astle et al., 2016.^65^

### ACO association signals

In stage 1, there were 83 distinct signals at P<5×10^−6^ (see **Figure 2, Supplementary Note** and **Supplementary Figure 2** for the signal selection process, and **Supplementary Table 5** for results). Of these, 31 signals retained significance (P<0.01) in further analyses comparing ACO cases separately with either COPD cases or asthma cases to determine whether signals were being driven by asthma or COPD alone (passes_prioritisation=TRUE in **Supplementary Table 5**). These were followed up in independent cohorts (stage 2). In stage 2, comprising 12 independent cohorts with an additional 4301 cases and 48609 controls (**Supplementary Table 1** and **Supplementary Table 2**) of European ancestry, 26/31 signals had a direction of effect concordant with the stage 1 discovery analysis (**Supplementary Table 6**). Whilst the sample size of individuals of African-American ancestry was small (297 cases, 1335 controls from the COPDGene study) and the confidence intervals around point estimates broad, 22/31 signals had a consistent direction of effect (**Supplementary Table 6)**.

**Figure 2.**
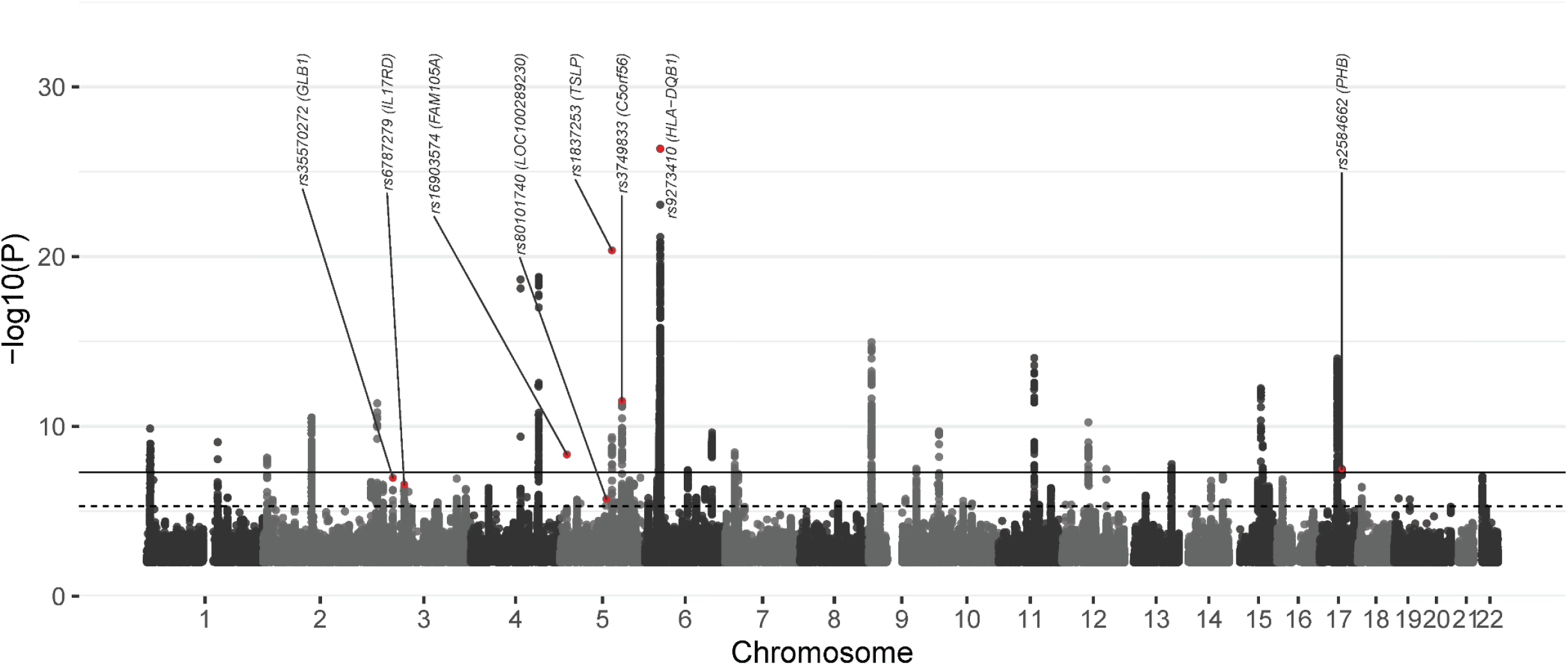
Manhattan plot of association results for ACO in stage 1 (UK Biobank). The x-axis shows genomic location by chromosome, the y-axis shows the –log_10_ P-value, corrected for the intercept of LD score regression (1.018). Eight top signals (from joint analysis) are highlighted in red, and labelled with rsIDs. The black line indicates p=5×10-8 (commonly known as genome-wide significance), and the dotted line corresponds to p=5×10-6 (genome-wide suggestive threshold). A quantile-quantile plot in shown in **Supplementary Figure 2**. For further details on the eight SNPs shown here, see also **Table 2**.

Results for a stage 2 sensitivity analysis including up to 9638 cases and 128273 controls of European ancestry from 15 studies, where COPD was defined either by available spirometry or, alternatively, by diagnoses from electronic health records (see **Supplementary Note**), are in **Supplementary Table 7**).

### Subgroup analyses

We repeated association testing in European ancestry individuals in stage 1 (UK Biobank) at the sentinel SNPs for the 31 signals entering stage 2 analysis after dividing the ACO cases into those with child- and adult-onset asthma (**Supplementary Table 8, Supplementary Figure 3**). Effect sizes amongst cases with childhood-onset asthma were highly correlated (and similar in magnitude) with those amongst individuals with onset in adulthood (R = 0.883). We also repeated association testing at these 31 sentinel SNPs after stratifying the sample by ever- and never-smoking status (**Supplementary Table 8** and **Supplementary Figure 4**). Again, effect sizes in ever- and never-smokers were closely correlated (R = 0.911), and similar in magnitude.

### Eight top signals from joint analysis

After meta-analysis combining stage 1 (UK Biobank) and stage 2 (12 independent cohorts) results in European ancestry individuals for each of the 31 sentinel SNPs followed up, 13 were genome-wide significant (p<5×10-8) (**Supplementary Table 5**; **Supplementary Figure 2** for flow diagram). Of these, eight either had a lower p-value in the joint analysis than in stage 1 (UK Biobank) alone, or had p<0.05 in the follow-up studies alone (**Table 2**, region plots in **Supplementary Figure 5**). None of these eight signals had been previously reported as being associated specifically with ACO.^8^ There are no reports of association with asthma, COPD or lung function for the intergenic signal on chromosome 5 (rs80101740, nearest gene *LOC100289230*, **Table 2**). Two signals, rs9273410 at the *HLA-DQB1* locus, and rs3749833 at the *C5orf56* locus, have been separately reported as associated with both asthma and lung function (and COPD in the case of rs9273410).^30, 33, 47, 52^ Intronic variant rs6787279 in *IL17RD* was previously reported to be associated with COPD and lung function but not with asthma.^53, 56, 63^ Four signals were previously reported to be associated with asthma but not with COPD – these were intronic variant rs35570272 in *GLB1*,^33^ exonic rs16903574 in *FAM105A*,^33^ intergenic rs2584662 at the *PHB* locus,^33^ and intergenic rs1837253 at the *TSLP* locus.^24, 26, 30^

**Table 2.** Eight genome-wide signals for asthma-COPD overlap.

| rsid | chr:pos<br>(effect/non-effect<br>allele) | Nearest<br>gene | Location | EAF | Stage 1 (UK Biobank,<br>cases=8068,<br>controls=40360) |  | Stage 2 (12 independent<br>studies, cases=4301,<br>controls =48609) |  | Joint analysis of stage 1<br>and stage 2 |  |
| --- | --- | --- | --- | --- | --- | --- | --- | --- | --- | --- |
|  |  |  |  |  | OR<br>(95% CI) | P | OR<br>(95% CI) | P | OR (95% CI) | P |
| rs35570272 | 3:33047662 (T/G) | *GLB1* | intronic | 0.398 | 1.11<br>(1.07, 1.15) | 1.06E-07 | 1.08<br>(1.02, 1.14) | 4.67E-03 | 1.10<br>(1.06, 1.13) | 2.44E-09 |
| rs6787279 | 3:57163751 (C/T) | *IL17RD* | intronic | 0.169 | 0.88<br>(0.84, 0.92) | 2.69E-07 | 0.91<br>(0.85, 0.97) | 6.51E-03 | 0.89<br>(0.86, 0.93) | 7.87E-09 |
| rs16903574 | 5:14610309 (G/C) | *FAM105A* | exonic | 0.077 | 1.23<br>(1.15, 1.32) | 4.47E-09 | 1.13<br>(1.03, 1.25) | 9.96E-03 | 1.20<br>(1.13, 1.27) | 3.8E-10 |
| rs80101740 | 5:98471135 (C/A) | *LOC1002892*<br>*30* | intergenic | 0.015 | 1.44<br>(1.24, 1.68)* | 1.87E-06* | 1.37<br>(1.10, 1.71) | 5.49E-03 | 1.42<br>(1.25, 1.61) | 3.72E-08 |
| rs1837253 | 5:110401872 (C/T) | *TSLP* | intergenic | 0.739 | 1.22<br>(1.17, 1.27) | 4.22E-21 | 1.06<br>(1.00, 1.12) | 4.44E-02 | 1.16<br>(1.12, 1.20) | 1.53E-18 |
| rs3749833 | 5:131799626 (C/T) | *C5orf56* | ncRNA_intronic | 0.263 | 1.16<br>(1.11, 1.21) | 3.10E-12 | 1.06<br>(1.00, 1.12) | 4.21E-02 | 1.12<br>(1.09, 1.16) | 9.37E-12 |
| rs9273410 | 6:32627250 (A/C) | *HLA-DQB1* | UTR3 | 0.445 | 1.24<br>(1.19, 1.29) | 4.37E-27 | 1.11<br>(1.05, 1.18) | 6.42E-04 | 1.20<br>(1.16, 1.24) | 9.19E-28 |
| rs2584662 | 17:47470487 (C/A) | *PHB* | intergenic | 0.42 | 0.90<br>(0.86, 0.94) | 3.20E-08 | 0.95<br>(0.90, 1.00) | 5.89E-02 | 0.92<br>(0.89, 0.95) | 2.21E-08 |

Variants were annotated with nearest gene and type of region using ANNOVAR software (and genome build hg19).<sup>78</sup> OR, 95% CI and P-value all calculated using score test. Firth test for rs80101740 gave OR 1.40 (95% CI 1.22, 1.60) and P=1.56E-06.

For each of the eight top signals, we performed fine-mapping to identify the most likely causal variants (the credible set), and sought associations with expression levels (eQTLs) and chromatin interactions. For the novel intergenic signal for ACO on chromosome 5 (rs80101740, EAF=0.015, OR 1.42, P=3.72×10^−8^) (see **Supplementary Table 6**), which has not been previously identified as associated with asthma, lung function or COPD, the sentinel SNP, rs80101740, had the largest posterior probability in the credible set (0.77), i.e. it had the largest probability of being the true causal variant, assuming the causal variant was genotyped or imputed. There was no evidence of colocalisation with eQTL signals at the rs80101740 locus (**Supplementary Table 11**), and no chromatin interactions were identified.

Four of our novel signals for ACO were previously reported for asthma but not COPD or lung function.^24, 30, 33^ rs35570272 in *GLB1* was associated with ACO with an odds ratio of 1.10 (EAF 0.398, P=2.44×10^−9^). There were 11 SNPs in the credible set, of which the intronic sentinel SNP had the highest posterior probability (0.655), i.e. was most likely to be the source of the association signal. There were significant chromatin interactions nearby in *GLB1* in fetal lung fibroblasts. *GLB1* encodes the beta-galactosidase enzyme involved in lysosomal function, and via alternative splicing also encodes an elastin-binding protein involved in the formation of extracellular elastic fibres. *GLB1* has been implicated in mucopolysaccharidosis type IV and GM1 gangliosidosis.^66^ Two SNPs (both with a posterior probability around 0.13) in the 99% credible set for this signal, rs7646283 and rs34064757 were eQTLs for cartilage associated protein (*CRTAP*) in lung (**Supplementary Table 12**), involved in bone development and implicated in osteogenesis imperfecta (type 7).

Another signal (previously reported for asthma) was rs16903574 (EAF=0.077, OR=1.20, P=3.8×10^−10^), a missense variant in *FAM105A*, which was identified as deleterious according to its CADD score (22.6), and predicted to result in an amino acid change from phenylalanine to leucine.^67^ *FAM105A* encodes a pseudoenzyme of unclear function, but possibly involved in protein-protein interactions.^68^ This sentinel SNP also had a posterior probability of 0.99, suggesting it is highly likely to be the true causal variant, assuming the causal variant was genotyped or imputed. A previous study in asthma also suggested *FAM105A* was the predicted target based on chromatin interactions and correlation between enhancer epigenetic marks and gene expression levels, though we did not identify any eQTL evidence in lung or whole blood.^33^ We also identified a highly significant chromatin interaction in fetal lung fibroblasts overlapping *FAM105A* and another nearby gene (*TRIO*), but not in adult lung.

An intergenic signal we identified between *PHB* and *ZNF652* (rs2584662; EAF=0.42, OR=0.92, P=2.21×10^−8^) has previously been associated with asthma and reported as a strong eQTL for *GNGT2* (implicated in NF-κB activation) in blood,^33, 34^ though we did not identify this in our lung and whole blood eQTL analysis. In our analysis there were eight SNPs in the credible set, none of which had a posterior probability over 0.2. Hi-C data suggested a significant chromatin interaction in *ZNF652*, with another less significant peak close to *GNGT2*. Nearby loci in *ZNF652* have previously been associated with asthma/allergic disease and moderate-to-severe asthma.^33^

We also identified rs1837253, an intergenic signal near *TSLP* (EAF 0.739, OR 1.16, P=1.53×10^−18^), with a posterior probability of 1, i.e. it was the only variant in the credible set. No eQTL evidence was identified, but there were highly significant chromatin interactions in fetal lung fibroblasts in *SLC25A46* and between *TSLP* and *SLC25A46*. The latter gene has been implicated in HMSN/Charcot-Marie-Tooth type 6, whilst the cytokine TSLP was implicated in asthma and allergic disease prior to the GWAS era,^69^ and an anti-TSLP antibody has been trialled in allergic asthma.^70^

Another signal, rs6787279 in *IL17RD*, has previously been associated with lung function and COPD (EAF 0.169, OR 0.89, P=7.87×10^−9^).^53, 56^ There were 55 variants in the corresponding 99% credible set, and the highest posterior probability of any variant was 0.12, meaning it is not yet possible to fine-map this signal with high confidence. One SNP in the credible set (rs17057718) was exonic and predicted to result in a possibly damaging amino acid change from valine to methionine, but the posterior probability was only 0.012. Three other SNPs were in the UTR3, but were not deleterious according to their CADD score (0). Multiple SNPs at this locus were eQTLs for *IL17RD* in lung, with the ACO risk increasing allele corresponding to decreased *IL17RD* expression. IL17RD is a component of the IL17 receptor complex; IL17 signalling has been implicated in the pathogenesis of COPD,^71, 72^ potentially by mediating the effects of cigarette smoke, as well as in asthma.^73^

Two ACO signals have previously been reported separately for both asthma and lung function or COPD: rs9273410 in *HLA-DQB1* (ACO OR 1.20, EAF 0.445, P=9.19×10^−28^) and rs3749833 in *C5orf56* (ACO OR 1.12, EAF 0.263, P=9.37×10^−12^). *HLA-DQB1* encodes a major histocompatibility complex (MHC) type II molecule involved in antigen presentation. Certain *HLA-DQB1* alleles have been associated with a range of inflammatory and autoimmune diseases, including type I diabetes and coeliac disease. In our analysis, the sentinel was the only SNP in the credible set. This was a UTR3 variant, but not deleterious according to its CADD score (0). For lung function, a specific amino acid change (from non-alanine to alanine) in the gene product HLA-DQβ1 has been identified as the main driver of signals in the MHC region.^63^ Analyses in asthma have identified *HLA-DQA1* as the likely driver gene.^33^

*C5orf56* is located on an important cytokine gene cluster on chromosome 5, which includes *IL3, IL4* and *IL5*. A number of the interleukins encoded in this region have been considered as possible therapeutic targets in asthma. IL5 in particular is now an important drug target in severe eosinophilic asthma, in which the anti-IL5 monoclonal antibodies mepolizumab and reslizumab have been shown to reduce exacerbation rates and improve quality of life in patients with severe eosinophilic asthma.^74-76^ SNPs in the credible set at this locus were eQTLs for *SLC22A5* in lung and blood, *AC116366*.6 in blood, *RAD50* in lung and a non-coding Y RNA in lung and blood. *SLC22A4* has previously been identified as the most likely candidate gene for the association with lung function.^63^ The gene products of both *SLC22A4* and *SLC22A5* are membrane transport proteins involved in bronchial uptake of bronchodilators (SLC22A5) and anti-cholinergic drugs used in asthma and COPD (SLC22A4/SLC22A5).^77^ An analysis in asthma predicted *C5orf56* (which encodes the interferon regulatory factor 1 (IRF1) antisense RNA) as the causal gene, based on enhancer-promoter chromatin interactions and a significant correlation between enhancer epigenetic marks and gene expression levels.^33^

We used PhenoScanner to undertake a phenome-wide scan for other associations with SNPs in each credible set, at FDR<0.01 (Supplementary Table 12). All of the ACO loci previously associated with asthma showed association with blood cell counts, particularly eosinophils and neutrophils, and atopic traits. The locus in the HLA region was associated, as expected, with a wide range of autoimmune and inflammatory traits. Another locus (rs2584662, near *PHB* and *ZNF652*), was associated with height, anthropometric traits, hypertension, mitral valve disease, coronary artery disease and traits indicating chronic disease and multimorbidity, whilst rs3749833 (near *C5orf56*), was associated with height and other anthropometric traits, and inflammatory bowel disease. SNPs in the credible set for the intergenic chromosome 5 signal (sentinel rs80101740), were associated with death from peripheral vascular disease and aortic stenosis, cervical cancer, postprocedural disorders of eye and adnexa, vaginal/uterine prolapse and phenoxymethylpenicillin treatment.

## Discussion

We conducted the largest GWAS of ACO to date, and identified 83 distinct signals associated with ACO at P<5×10^−6^ in stage 1. After excluding variants associated with asthma only or COPD only, we studied 31 variants in another 12 studies and discovered eight distinct signals for ACO that showed genome-wide significance in a meta-analysis of stage 1 and stage 2 studies.

Our study contributes to understanding of the genetic architecture of ACO. As measured by genome-wide genetic correlation, we showed strong genetic overlap between ACO and COPD, ACO and severe asthma, and ACO and asthma. Furthermore, we found genetic correlation between ACO and blood eosinophil counts. Increased eosinophil levels have been associated with exacerbations in asthma and COPD,^79-81^ and with decline in lung function in asthmatic and non-asthmatic subjects.^82^ We also noted that eosinophil counts, atopy and asthma traits were prominent when we ran phenome-wide scans of our top eight signals. These findings are consistent with an important role for type 2 inflammation in ACO.^83, 84^

We identified an intergenic signal on chromosome 5, rs80101740, which had not previously been associated with asthma, COPD or lung function. Whilst this signal is near to a putative signal for lung function without replication support (rs377731, r^2^=0.02 with rs80101740),^63^ the lead ACO sentinel persisted after conditioning on the lung function signal. Evidence from eQTL studies suggests that the nearby lung function signal is associated with expression of *RGMB* and *LINC02062*.

Four of the eight signals we identify as novel for ACO (*GLB1, FAM105A, PHB, TSLP*) are known signals for asthma or allergic disease but not COPD. Our results suggest that these loci also have a role in fixed airflow obstruction. All four of these signals have been associated with both child- and adult-onset asthma, and therefore could represent an opportunity to intervene in early life to prevent serious long-term sequelae.^38^ One ACO signal (*IL17RD*) is a known locus for lung function and COPD, and our findings demonstrate the relevance of this locus in reversible airflow obstruction. Taken together, these loci could represent targets for intervention, potentially to prevent development of fixed airflow obstruction.

Two signals had previously been reported as associated with asthma and either COPD or lung function, including the *HLA-DQB1* locus, the first to be identified as being associated with both asthma and COPD, and a signal at *C5orf56*, encoding interferon regulatory factor I (*IRF1*) antisense RNA, located on chromosome 5 near a cytokine gene cluster.

We undertook subgroup analyses to examine whether smoking status or age of asthma onset influenced effect size estimates at 31 association signals from stage 1. Across these variants, there was a strong positive correlation between the effect sizes for ACO in ever- and never-smokers, showing that overall, being an ever-smoker was not driving the associations identified. This suggests that ACO is not due solely to smoking in people with asthma, although it is known that childhood asthma in smokers increases risk of COPD in later life compared with non-asthmatics, possibly due to reduced lung growth in early life.^35^ Similarly, when cases were stratified by whether the asthma-component of ACO was diagnosed in childhood or adulthood, there was a strong correlation between effect sizes in both groups.

There are a number of potential limitations to our study. The follow-up sample size (4301 cases) was substantial, although relatively underpowered compared to the discovery (8068 cases). Misclassification of asthma and COPD diagnoses is possible: for example, the presentation of asthma in older patients may mimic COPD, and clinicians are less likely to suspect COPD in patients who do not smoke. To mitigate this, we utilised GOLD 2-4 spirometric criteria (FEV_1_/FVC<0.7 and FEV_1_ <80% predicted) to define COPD wherever possible. Self-reported asthma has previously been validated in populations at risk of COPD.^35^ Any remaining misclassification would have attenuated effect estimates towards the null, that is, reduced the power to detect true genetic associations with ACO. Our main analysis was undertaken in European ancestry populations only; although for many loci there was good concordance in a small sample of African-American ancestry from the COPDGene study, it is essential to study this trait further in diverse populations.

In the largest genome-wide association study of asthma-COPD overlap to date, we identified eight signals associated with ACO. Our findings suggest a spectrum of shared genetic influences from variants which predominantly influence asthma, to those which predominantly influence fixed airflow obstruction. We focus here on variants that tend towards an intermediate phenotype with features of both asthma and fixed airflow obstruction, with pathways implicating innate and adaptive immunity and potentially bone development, as well as signals for which the biology is as yet unclear. Further understanding of the biology of these signals is likely to be important for therapeutics to prevent the development of fixed airflow obstruction among people with asthma.

## Methods

### Stage 1

#### Data source and study population

The data source for this study was UK Biobank (http://www.ukbiobank.ac.uk).

Individuals were eligible for inclusion in this study if they met the following criteria: (i) had data on age, sex and height; (ii) had spirometry that met quality control requirements (acceptability, reproducibility and blow curve metrics); (iii) had genome-wide imputed genetic data that met quality control requirements; and (iv) were of European ancestry based on *k*-means clustering after principal components analysis. Quality control processes have been described previously.^63^

Genotyping was undertaken using the Affymetrix Axiom® UK BiLEVE array and the Affymetrix Axiom® UK Biobank array,^47^ with imputation to the Haplotype Reference Consortium panel.^85^ 321,057 individuals were eligible for inclusion in this analysis and 37 million SNPs were available for analysis.

#### Case and control selection

Individuals were selected as cases of ACO if they had evidence of asthma from either the touchscreen questionnaire (UK Biobank data field 6152) or verbal interview (field 20002) AND FEV_1_/FVC <0.7 with classification of airflow limitation GOLD 2+ (FEV_1_ <80% of predicted) at any study visit. Individuals were excluded from the cases if they reported a diagnosis of alpha-1-antitrypsin deficiency in the verbal interview (field 20002) at any study visit. Where there were related pairs within the cases (second degree or closer), the individual with the lower genotype call rate was excluded.

Controls were free of both asthma and COPD, based on no reported bronchitis, emphysema or asthma on the touchscreen questionnaire, and no reported asthma, COPD, emphysema/chronic bronchitis, bronchitis, emphysema, alpha-1-antitrypsin deficiency on the verbal interview. Controls had FEV_1_ ≥80% predicted and FEV_1_/FVC >0.7. Individuals related to cases were excluded, and where pairs of controls were related, the individual with the lower genotype call rate was excluded (as for cases). Controls were randomly selected in a ratio of five to each case.

Two additional control sets were defined for the purpose of signal prioritisation: individuals with evidence of asthma but without COPD, and individuals with evidence of COPD but without asthma. For these analyses, asthma and COPD were defined as described above. Related individuals were excluded from these control sets as described above.

#### Genome-wide analysis

Association testing was undertaken using the ‘score’ option implemented in SNPTEST (https://mathgen.stats.ox.ac.uk/genetics_software/snptest/snptest.html, version 2.5.2), under an additive model of genotype dosage. Age, sex, smoking status (ever/never smoking), genotyping array and the first 10 principal components were included as covariates. Variants were filtered based on a minor allele frequency (MAF) greater than 0.01 and imputation quality (info) greater than 0.5.

The genomic inflation factor (λ) was calculated as the intercept from LD score regression (LDSC, https://github.com/bulik/ldsc), which should be robust to inflation by polygenicity. P-values and standard errors were recalculated from χ^2^ statistics adjusted for λ.

Sentinel variants and regions of association were identified by taking the variant with the lowest P-value and extracting this sentinel variant and the surrounding region ±1Mb. Then, the variant with the next lowest P-value outside this region was identified, proceeding iteratively until there were no further variants with a P-value less than 5×10^−6^. All coordinates are given according to GRCh37.

To identify distinct signals, and additional signals within the regions described above, conditional analyses were undertaken using GCTA-COJO (http://cnsgenomics.com/software/gcta/#COJO). For non-HLA sentinel variants, GCTA-COJO was used to implement a joint, stepwise selection procedure for each of the sentinels and its corresponding 2Mb surrounding region. For the HLA region, one conditional analysis was performed for the region of chr6:26,000,000-34,000,000.

After the set of distinct signals were identified, two further analyses were undertaken in order to ascertain the extent to which signals were associated with COPD and/or asthma, for signal prioritisation. These analyses included the same cases as the primary analysis, plus the two control sets described above: i) controls with COPD but no asthma and ii) controls with asthma but no COPD. Analyses for conditional-only signals (e.g. signals where P<5×10^−6^ in conditional analyses only) were run in SNPTEST, by conditioning on the sentinel variant.

Variants were selected for follow-up in stage 2 if they were identified as distinct signals in the main stage 1 analysis (P<5×10^−6^ for association with ACO), and were also associated with ACO at P<0.01 in both signal prioritisation analyses.

#### Genetic correlation analysis

Using LD score regression,^64^ we computed genetic correlations between ACO, asthma, and FEV_1_/FVC. We used ACO GWAS results from the current analysis (stage 1), results from a recent GWAS of asthma (child- and adult-onset),^34^ an additional GWAS of moderate-severe asthma,^55^ a GWAS of COPD,^56^ and FEV_1_/FVC GWAS results from our genome-wide meta-analysis of UK Biobank and the SpiroMeta consortium.^63^ We used LD-Hub to compute genetic correlations between ACO and other atopic, auto-immune, and smoking behaviour traits (http://ldsc.broadinstitute.org/).^86^

### Stage 2

#### Studies and meta-analysis

SNPs identified in stage 1 were tested for association in twelve independent studies of European ancestry populations (up to a total of 4,301 cases and 48,609 controls, in CHS, COPDGene, deCODE, ECLIPSE, EPIC-Norfolk, FHS, Generation Scotland, HUNT, LOVELACE, Mass General Brigham Biobank, Rotterdam Study, SPIROMICS) and one African-American ancestry cohort from the COPDGene study (297 cases, 1335 controls). See the **Supplementary Note** for study descriptions, **Supplementary Table 1** for phenotype information on each study, and **Supplementary Table 2** for information on the genotyping and imputation.

Cases were required to have both asthma and COPD. Asthma was defined as any self-report of asthma in the individual’s lifetime (most studies specified self-report of *doctor-diagnosed* asthma), or an asthma diagnosis in the healthcare record (including billing codes). We required cases to have spirometry indicating COPD with GOLD2+ airflow obstruction (FEV_1_/FVC<0.7, and FEV_1_<80% predicted). Controls had normal spirometry (FEV_1_/FVC>0.7, and FEV_1_≥80% predicted) and no asthma diagnosis. Where possible, studies excluded individuals with alpha-1-antitrypsin deficiency on the basis of clinical or genetic data.

Studies undertook logistic regression with ACO as the outcome, using an additive genetic model, and adjusting for age, sex and, where available, smoking status. An appropriate number of ancestry principal components were also included, or a mixed linear model used to account for fine-scale population structure or familial relatedness. Proxies (r^2^>0.3) were provided if SNPs were missing, or if a SNP was poorly imputed (info <0.5) in a study (see **Supplementary Table 3**). Results were combined across stage 2 studies using fixed-effect meta-analysis, using the ‘meta’ package of R. Heterogeneity was assessed using the I^2^ statistic. We then combined these results with those from UK Biobank (stage 1).

We additionally performed a sensitivity analysis incorporating all cohorts included in the main analysis, but expanding the definition of COPD to include a diagnosis in the healthcare record (including billing codes) in three cohorts where this information was available (deCODE, HUNT, Rotterdam) and three additional cohorts (FinnCAD, GenKOLS, GERA). COPD controls were defined based on normal spirometry, and/or absence of a diagnosis in the healthcare record (see **Supplementary Note**).

To focus bioinformatic analyses on signals with the strongest evidence of association with ACO, we first focused on to those that were genome-wide significant (p<5×10-8) in the joint analysis of stage 1 and stage 2. We then restricted this group of signals to those that had a lower p-value in the joint analysis than in UK Biobank (stage 1) alone, or had p<0.05 in stage 2.

#### Subgroup analyses

To assess whether the associations at our signals identified in stage 1 changed according to the age at which cases were diagnosed with asthma, we divided cases into those reporting an asthma diagnosis in childhood (<12 years), and those reporting an asthma diagnosis in adulthood (>25 years), and repeated the association tests in UK Biobank.^38^ The age thresholds were chosen in order to minimise the risk of misclassification, following an approach taken in other GWAS of childhood and adult-onset asthma. In addition, we repeated association testing at the same stage 1 signals after stratifying our sample by ever- and never-smoking status.

### Bioinformatic analyses on top signals

#### Signal fine-mapping and functional annotation

For each of the top signals (defined as described above), we identified the set of SNPs that was 99% likely to contain the causal variant (assuming that the causal variant was included in the dataset), as described previously.^63, 87^

We used wANNOVAR (http://wannovar.wglab.org/) to annotate variants in the credible sets. We then used SIFT,^88^ FATHMM [implemented using https://www.ensembl.org/vep and http://fathmm.biocompute.org.uk/],^89^ and PolyPhen-2 [implemented using the HumDiv model via http://genetics.bwh.harvard.edu/pph2/]^67^ to annotate exonic variants, and CADD to additionally annotate UTR variants.^63, 90^

#### Association of ACO signals with gene expression (eQTL) and chromatin interactions

We queried the 99% credible sets for the top signals against expression quantitative trait locus (eQTL) resources in lung tissue (n=1,038),^47, 91-93^ and the GTEx online portal (v8) [lung N=515, whole blood N=670],^94^ in order to determine whether the locus was significantly associated with gene expression, i.e. whether it was an eQTL.

Functional information about nearby genes, or those implicated through eQTL analysis, was retrieved from the National Institute of Health Genetics Home Reference.

For one signal which has not previously been associated with asthma, lung function or COPD, we additionally performed formal testing for Bayesian colocalisation with cis-eQTL signals (gene genomic position ± 0.5 Mb) using the R package COLOC.^95^ The method has been described previously,^96^ briefly tests whether GWAS signal and eQTL signal for this region are consistent with shared causal variant(s). A large posterior probability is supporting evidence of a single shared causal variant for the two traits.

We additionally entered the sentinel SNP for each signal into the Hi-C Unifying Genomic Interrogator (HUGIn) and looked for long-range chromatin interactions with other genomic regions (in adult lung and fetal lung fibroblasts) where the false discovery rate (FDR) was less than 5%.^97-99^

#### Association of ACO signals with other phenotypes

To assess the associations of SNPs in the credible sets with other traits, we undertook a phenome-wide scan using PhenoScanner (http://www.phenoscanner.medschl.cam.ac.uk/).^100^ We calculated FDRs using the Benjamini-Hochberg method and used a threshold of FDR less than 1%. Evidence of association with rare diseases was also sought in OMIM and Orphanet for the nearest gene to any SNP in the credible set, as well as genes implicated by eQTL evidence.

## Data Availability

Summary statistics are available from the authors on request and will be made available via public portals upon publication.

## Acknowledgements

The research was conducted using the UK Biobank resource (http://www.ukbiobank.ac.uk), under application 648.

This work is supported by BREATHE, the Health Data Research Hub for Respiratory Health [MC_PC_19004] in partnership with SAIL Databank. BREATHE is funded through the UK Research and Innovation Industrial Strategy Challenge Fund and delivered through Health Data Research UK.

Please see the supplementary materials for additional study acknowledgements and funding statements.

## Author Contributions

C.J., A.L.G., L.V.W., M.D.T. contributed to the conception and design of the study. C.J., A.L.G., N.S., T.A.O., J. Liu, L.P.H., S.H.C., J.K., J. Luan, X.L., N.T., H.X., T.M.B., H.P., S.L., A.C., A.I.H.C., M.O. undertook data analysis. C.J., A.L.G., N.S., R.P., S.A.B. M.O., G.T., D.A.M., E.R.B., L.C.S., C.I., Y.T., S.A.G., J.D., L.L., V.E.O., I.J., J.K.Q., D.D.S., Y.B., M.v.d.B, D.C.N., I.S., I.P.H., C.L., S.R., T.L., A.C.W., J. Lasky-Su, P.B., A.G., C.P.H., C.H., A.L., B.B., K.S., M.H.C., L.V.W., M.D.T. contributed to data acquisition and/or interpretation. C.J., A.L.G., L.V.W., M.D.T. drafted the manuscript. All authors critically reviewed the manuscript before submission.

## Conflicts of Interest

T.A.O., G.T., I.J. and K.S. are employees of deCODE genetics/Amgen Inc. E.R.B. has undertaken clinical trials through his employer, Wake Forest School of Medicine and University of Arizona, for AstraZeneca, MedImmune, Boehringer Ingelheim, Genentech, Novartis, Regeneron, and Sanofi Genzyme. E.R.B. has also served as a paid consultant for ALK-Abello, AztraZeneca, Glaxo Smith Kline, MedImmune, Novartis, Regeneron, Sanofi Genzyme, and TEVA. M.v.d.B. reports grants paid to the University from Astra Zeneca, TEVA, GSK and Chiesi outside the submitted work. D.C.N. has been a Merck & Co. employee during this study and is now an employee at Biogen Inc. I.S. has received support from GSK and Boehringer Ingelheim. I.P.H. has funded research collaborations with GSK, Boehringer Ingelheim and Orion. M.H.C. has received grant support from GSK and Bayer, and consulting and speaking fees from Genentech, Illumina and Astrazeneca. L.V.W. receives funding from GSK for a collaborative research project outside of the submitted work. M.D.T. receives funding from GSK and Orion for collaborative research projects outside of the submitted work.

## Funding

C.J. holds a Medical Research Council Clinical Research Training Fellowship (MR/P00167X/1). A.L.G. was funded by internal fellowships at the University of Leicester from the Wellcome Trust Institutional Strategic Support Fund (204801/Z/16/Z) and the BHF Accelerator Award (AA/18/3/34220). L.P.H. is funded by NIH/NHLBI (5K23HL136851). S.H.C. is funded by NIH/NHLBI (1K01HL153941-01). A.I.H.C. is supported by MITACS accelerate. M.H.C. is supported by R01 HL137927, R01 HL089856, and R01 HL147148. L.V.W. holds a GSK/British Lung Foundation Chair in Respiratory Research. M.D.T. is supported by a Wellcome Trust Investigator Award (WT202849/Z/16/Z). M.D.T. and L.V.W. have been supported by the MRC (MR/N011317/1). M.D.T. and I.P.H. hold NIHR Senior Investigator Awards.

The research was partially supported by the NIHR Leicester Biomedical Research Centre and the NIHR Nottingham Biomedical Research Centre; the views expressed are those of the author(s) and not necessarily those of the NHS, the NIHR or the Department of Health. The funders had no role in the design of the analyses. C.J. and M.D.T. were involved in all stages of study development and delivery, and M.D.T. had full access to all data in the study and final responsibility for the decision to submit for publication. Please see the supplementary materials for additional study acknowledgements and funding statements.

## Notes

### Author Declarations

The research was conducted using the UK Biobank resource (http://www.ukbiobank.ac.uk), under application 648. UK Biobank has Research Ethics Committee (REC) approval (11/NW/0382).

