## Supplementary Note and Figures for "Genetic associations and architecture of asthma-chronic obstructive pulmonary disease overlap"

### Supplementary Material

***For Supplementary Tables, see separate Excel file.***

#### Contents

Supplementary Note ... 2
Study Descriptions ... 2
Signal selection ... 4
Follow-up and meta-analysis ... 5
Supplementary Figure 1: Quantile-quantile plot of ACO GWAS results from discovery analysis in UK Biobank ... 6
Supplementary Figure 2: Flowchart detailing the signal selection process in the UK Biobank discovery analysis ... 7
Supplementary Figure 3 Scatterplots comparing effects in 31 signals taken forward for replication, with results when ACO cases are split into those with adult- (>25 years) and child-onset (<12 years) asthma ... 8
Supplementary Figure 4 Scatterplots comparing effects in 31 signals taken forward for replication, with smoking-stratified results (by ever- and never-smokers) ... 9
Supplementary Figure 5 Region plots for eight signals taken forward for downstream analyses. ... 10
rs80101740 ... 10
rs35570272 ... 10
rs16903574 ... 11
rs2584662 ... 11
rs1837253 ... 11
rs6787279 ... 12
rs9273410 ... 12
rs3749833 ... 12
Study acknowledgements and funding ... 13
References ... 16

### Supplementary Note

#### Study Descriptions

The **Cardiovascular Health Study (CHS)** is a population-based cohort study of risk factors for coronary heart disease and stroke in adults ≥65 years conducted across four field centers.[1] The original predominantly European ancestry cohort of 5,201 persons was recruited in 1989-1990 from random samples of the Medicare eligibility lists; subsequently, an additional predominantly African-American cohort of 687 persons was enrolled for a total sample of 5,888. Blood samples were drawn from all participants at their baseline examination and DNA was subsequently extracted from available samples. Genotyping was performed at the General Clinical Research Center's Phenotyping/Genotyping Laboratory at Cedars-Sinai among CHS participants who consented to genetic testing and had DNA available using the Illumina 370CNV BeadChip system (for European ancestry participants, in 2007) or the Illumina HumanOmni1-Quad_v1 BeadChip system (for African-American participants, in 2010). Only European ancestry participants were included in this analysis due to low numbers of African-Americans with the outcome. For European ancestry participants in this analysis, spirometry values to define COPD and self-reported asthma are from the 1989-1990 exam. CHS was approved by institutional review committees at each field center and individuals in the present analysis had available DNA and gave informed consent including consent to use of genetic information for the study of cardiovascular disease.

**COPDGene**: Eligible subjects in COPDGene Study (NCT00608764, www.copdgene.org) were of nonHispanic white (NHW) or African-American (AA) ancestry, aged 45-80 years old, with at least 10 packyears of smoking and no diagnosed lung disease other than COPD or asthma.[2,3] IRB approval was obtained at all study centers, and all study participants provided written informed consent. Illumina (San Diego, CA) performed genotyping on the HumanOmniExpress array.

**deCODE:** The Icelandic GWAS is based on 32.5 million variants identified through whole-genome sequencing of 28,075 Icelandic individuals and subsequently imputed for 155,250 Icelandic individuals, genotyped using the Illumina SNP chips, as well as 285,664 of their first- and second-degree relatives as previously described.[4]

**ECLIPSE:** Evaluation of COPD Longitudinally to Identify Predictive Surrogate End-points (ECLIPSE; SCO104960, NCT00292552, www.eclipse-copd.com): Details of the ECLIPSE study and genome-wide association 4 analysis have been described previously.[5] The ECLIPSE study was approved by the relevant ethics and review boards at the participating clinical centers. All participants provided written informed consent. Cases and controls were aged 40-75 with at least a 10 pack-year smoking history without other respiratory diseases. Genotyping was performed using the Illumina HumanHap 550 V3 (Illumina, San Diego, CA).

**European Prospective Investigation of Cancer (EPIC)-**Norfolk (DOI 10.22025/2019.10.105.00004) is a prospective population-based cohort study which recruited 25,639 men and women aged 40-79 years at baseline between 1993 and 1997 from 35 participating general practices in Norfolk, UK.[6] Individuals attended for a baseline health check including the provision of blood samples for concurrent and future analysis. Further health check visits have been conducted since the baseline visit. Participants have contributed information about their diet, lifestyle and health through questionnaires and health checks over two decades. DNA has been extracted from all EPIC participants and stored blood has been analysed for an extensive range of classical and novel biomarkers. Sample quality control was performed including gender check, relatedness check, and ancestry check. The Norwich Local Research Ethics Committee granted ethical approval for the study. All participants gave written informed consent.

**FinnCAD:** The Finnish Chronic Obstructive Airway Disease (CAD) cohort has been enrolled through the Pulmonary Clinics of Helsinki and Turku University Hospitals during the years 2005-2007.[7] All out- and inpatients aged 18–75 years who had been discharged with a diagnosis of asthma or COPD (ICD10 code J44–J46) were invited to the study and 1855 patients participated. Three hundred twenty one of the patients full filled both asthma and COPD criteria and were included to the study. Controls for this study were selected from the Generisk cohort.

**Framingham Heart Study** (FHS; NCT00005121): Details on pulmonary function in the FHS have been previously published.[8,9] FHS was IRB-approved at the relevant institutions, and all participants provided written informed consent. We analyzed data from the most recent exam for each of the three generations of families participating in the FHS were analyzed. Genotypes were from the Affymetrix 500K array supplemented by the Affymetrix MIPS 50K.

**Generation Scotland** is a multi-institution collaboration that has created an ethically sound, family-based and population-based resource for identifying the genetic basis of common complex diseases.[10] The Scottish Family Health Study component (GS:SFHS) has DNA and sociodemographic, psychological and clinical data from ~24,000 adult volunteers from across Scotland. The ethnicity of the cohort is 99% Caucasian, with 96% born in the UK and 87% in Scotland. Features of GS:SFHS include the family-based recruitment, breadth and depth of phenotype information, 'broad' consent from participants to use their data and samples for a wide range of medical research and for re-contact, and consent and mechanisms for linkage of all data to comprehensive routine healthcare records. These features were designed to maximise the power of the resource to identify, replicate or control for genetic factors associated with a wide spectrum of illnesses and risk factors

**GenKOLS** (Norway): The Norwegian GenKOLS (Genetics of Chronic Obstructive Lung Disease, GSK code RES11080) recruited subjects with > 2.5 pack years of smoking history from Bergen, Norway.[11] Subjects with severe alpha-1 antitrypsin deficiency and other lung diseases (aside from asthma) were excluded. The Regional Committee for Medical Research Ethics (REK Vest), the Norwegian Data Inspectorate and the Norwegian Department of Health approved the case–control study. Written informed consent was obtained from all participants. Genotyping was performed using Illumina HumanHap 550 arrays (Illumina, San Diego, CA).

**GERA**: This study utilized genome-wide genetic data available on the Genetic Epidemiology Resource in Adult Health and Aging (GERA) cohort of 110,266 adult male and female Kaiser Permanente of Northern California (KPNC) members.  The cohort has been described in detail elsewhere.[12] In brief, the GERA cohort was formed by including all racial and ethnic minority participants in the larger cohort of the Research Program on Genes, Environment and Health (RPGEH) with saliva samples (19% of the total); the remaining participants were drawn randomly from White non-Hispanic participants (81% of the total).  All RPGEH participants responded to a self-administered questionnaire in 2007/08 that included information on medical history, ancestry, health behaviors (smoking, alcohol consumption, diet, physical activity and reproductive history) and current weight and height.  Asthma and COPD phenotypes were derived from the KPNC electronic health record and linked with the genetic data by virtue of the unique medical record number.

**HUNT**: The Nord-Trøndelag Health Study (HUNT) is a population-based health survey conducted in the county of Nord Trøndelag, Norway. Individuals were included at four different time points during approximately 20 years (HUNT1 [1984-1986], HUNT2 [1995-1997], HUNT3 [2006-2008]) and HUNT4 [2017-2019])(PMID: 22879362). At each time point, the entire adult population (≥ 20 years) was invited to participate by completing questionnaires, attending clinical examinations and interviews. Participation rates have generally been high: 89.4% (n = 77,212), 69.5% (n = 65 237),

54.1% (n = 50 807) and 54.0% (n=56042) in HUNT1, HUNT2, HUNT3 and HUNT4, respectively(PMID: 22879362). Taken together, the health studies include information from over 120,000 different individuals from Nord-Trøndelag. Biological samples including DNA have been collected for approximately 90,000 participants.

**Lovelace**: The Lovelace Smokers Cohort (LSC) has been actively enrolling smokers from the Albuquerque, NM metropolitan area since 2001.[13] All participants provided written informed consent, and the study was approved by the relevant IRB. Enrollment was restricted to current and former smokers age 40 to 74 years old with a minimum of 10 pack-years of smoking and no personal history of lung cancer. A detailed questionnaire written in English was used to collect information on demographics; medical, cigarette smoking, and exposure history; socioeconomic status; diet; and quality of life. Pulmonary function testing was performed at each visit. All participants signed a consent form, and the Western Institutional Review Board approved this project. The GWAS discovery set was comprised of 1200 Caucasian (self-reported) smokers. The HumanOmni2.5-4v1-H BeadChip (Illumina, San Diego, CA) was used to genotype 2,450,000 SNPs in 1200 Caucasian smokers from the LSC.

The **Rotterdam Study** is a prospective population-based cohort study founded in 1990 in a suburb of Rotterdam, the Netherlands.[14,15] The first cohort (RS-I) consists of 7,983 participants, aged 55 years and over. The second cohort (RS-II) was recruited in 2000 with the same inclusion criteria. The third cohort (RS-III) consists of 3,932 participants, aged 45 years and over and was recruited in 2006. The Rotterdam Study was approved by the institutional review board (Medical Ethics Committee) of the Erasmus Medical Center and by the review board of The Netherlands Ministry of Health, Welfare and Sports. All participants provided written informed consent. Spirometry was performed using the Master Screen® PFT Pro (CareFusion, San Diego, CA). A total of 6,291 subjects for RS I, 2,157 for RS II and 3,048 for RS III passed genotyping quality control.

**SPIROMICS** is a prospective cohort (n=2,981) with the goal to identify COPD subphenotypes and biomarkers of disease progression.[16] SPIROMICS is a well-characterized longitudinal cohort with comprehensive phenotyping including measurements of lung function and quantitative CT scans. Smokers with COPD were defined as smokers (smoking≥20 packs/year) with post-bronchodilator FEV1/FVC<0.7 (GOLD stage 1-4) and 'healthy' smoking controls were defined as smokers (smoking≥20 packs/year) with post-bronchodilator FEV1/FVC≥0.7 (GOLD stage 0). Healthy non-smoking controls were also recruited. Participants were recruited at each center using a common detailed protocol (www.spiromics.com). The study was approved by the institutional review boards of all participating sites with written informed consent from all participants. DNA was isolated using standard protocols, SNP genotyping performed using Illumina HumanOmniExpressExome BeadChip and BeadStudio (Illumina, Inc., San Diego, CA), and quality control and genetic association analysis were described previously.[17]

#### Signal selection

There were 80 sentinel variants associated with ACO at $P<5x10^{-6}$, of which 28 reached genome-wide significance ($P<5x10^{-8}$). Of the 80 sentinels, four were located in the major histocompatibility complex (MHC) region (chr6:28,477,797-33,448,354), with a further three located just outside this region (chr6:26,000,000-34,000,000).

Conditional analysis of the 73 non-HLA sentinel variants identified five additional independent signals within 1Mb of these sentinels, plus two conditional signals ( only associated at $P<5x10^{-6}$ after conditioning on the sentinel variant), giving a total of 80 conditionally independent non-HLA signals. Separate conditional analysis of the extended HLA region identified three HLA signals.

#### Follow-up and meta-analysis

In addition to the main meta-analysis, we undertook a sensitivity meta-analysis in which we varied the definition of COPD. For most studies, this included the same cases as the main analysis, but we additionally allowed COPD cases to be defined by a diagnosis in the healthcare record (including billing codes), if that was available within the study. COPD controls were defined based on normal spirometry, and/or absence of a diagnosis in the healthcare record. As in the main analysis, studies undertook logistic regression with ACO as the outcome, using an additive genetic model, and adjusting for age, sex and, where available, smoking status. An appropriate number of ancestry principal components were also included, or a mixed linear model used to account for fine-scale population structure. Proxies ($r^2$>0.3) were provided if SNPs were missing or poorly imputed (info<0.5) in a study (see **Supplementary Table 3**).

9638 cases were included in this analysis, and 26/31 signals had a direction of effect that was concordant with the discovery analysis. The strongest signal was rs9273410, OR 1.103, 95% CI 1.055, 1.153, P=1.55x$10^{-5}$, $I^2$=0%, **Supplementary Table 7**). Of the 26/31 directionally concordant signals in each meta-analyses, 24/31 were directionally concordant in both analyses. Three of the 8 signals followed up in downstream analyses met the criteria for follow-up in this analysis, and no other signals of the remaining 23/31 met these criteria (p<5x10-8 in the joint analysis of stage 1 and stage 2, and either a lower p-value in the joint analysis than in UK Biobank alone, or p<0.05 in stage 2 alone).

#### Supplementary Figure 1: Quantile-quantile plot of ACO GWAS results from discovery analysis in UK Biobank

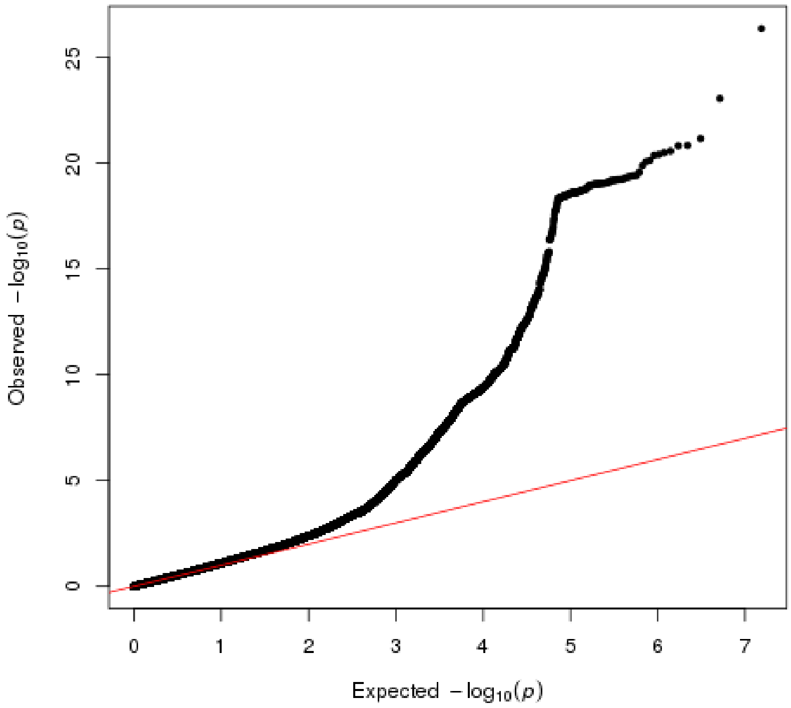


8068 ACO cases, 40360 controls, 7,693,381 SNPS. Intercept value from univariate LD score regression = 1.018. Standard errors and p-values in UK Biobank are corrected for this value.

#### Supplementary Figure 2: Flowchart detailing the signal selection process in the UK Biobank discovery analysis

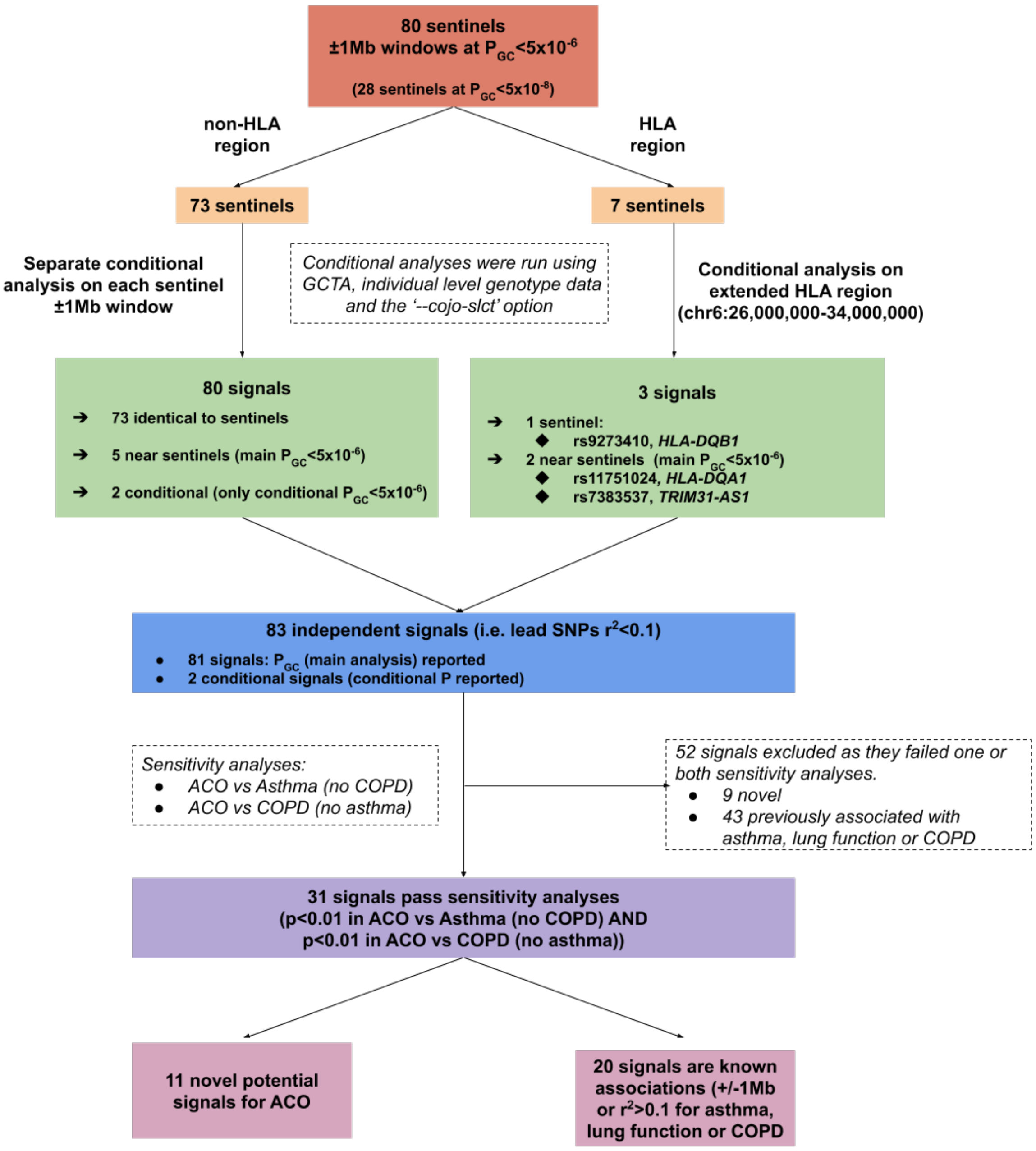

#### Supplementary Figure 3 Scatterplots comparing effects in 31 signals taken forward for replication, with results when ACO cases are split into those with adult- (>25 years) and child-onset (<12 years) asthma

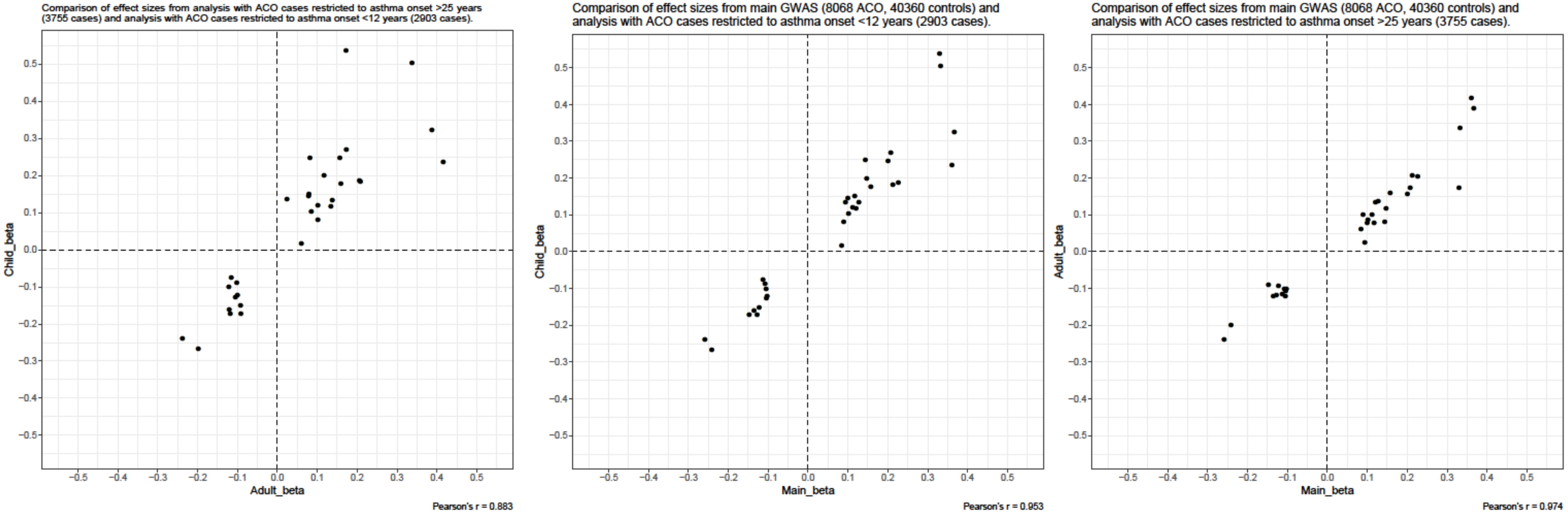


From left to right: comparison of effects (betas) of child-onset asthma ACO vs adult-onset asthma ACO cases; comparison of effects in child-onset ACO versus all ACO (e.g. main analysis); comparison of effects in adult-onset ACO versus all ACO (e.g. main analysis). Both subgroup analyses used the same 40,360 controls as used in the main analysis.

#### Supplementary Figure 4 Scatterplots comparing effects in 31 signals taken forward for replication, with smoking-stratified results (by ever- and never-smokers)

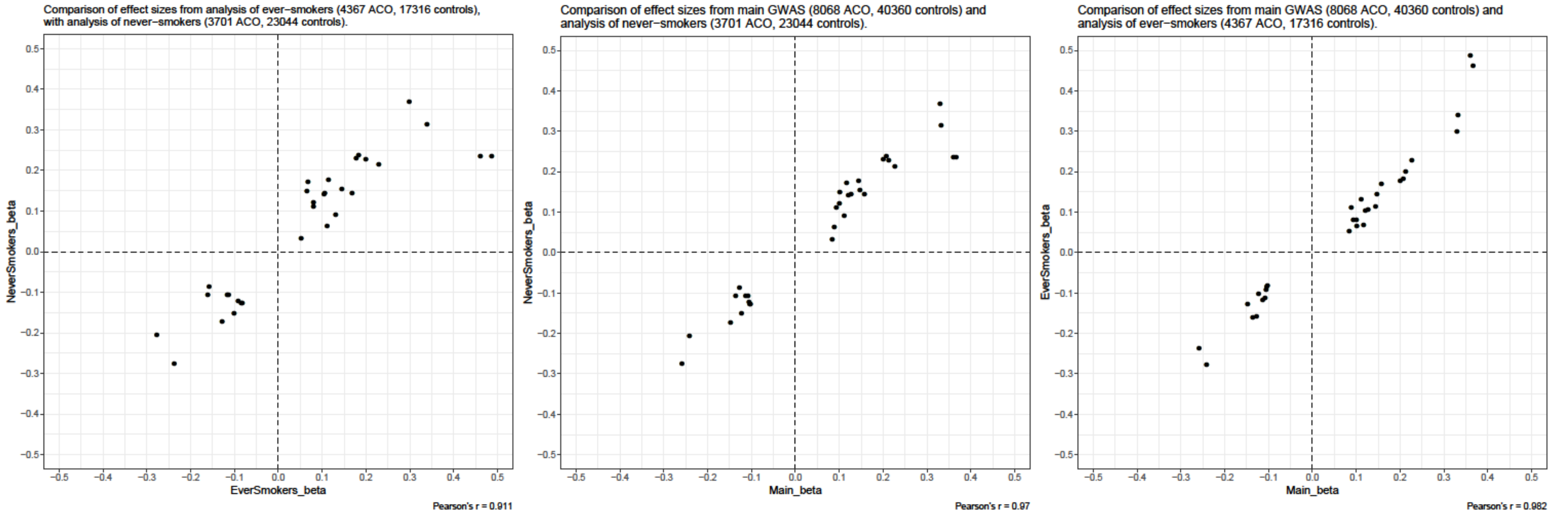


From left to right: comparison of effects (betas) of ACO versus controls (never-smokers) and ACO versus controls (ever-smokers); comparison of effects in ACO versus controls (never-smokers) versus all ACO (e.g. main analysis); comparison of effects in ACO versus controls (ever-smokers) versus all ACO (e.g. main analysis).

#### Supplementary Figure 5 Region plots for eight signals taken forward for downstream analyses.

### rs80101740

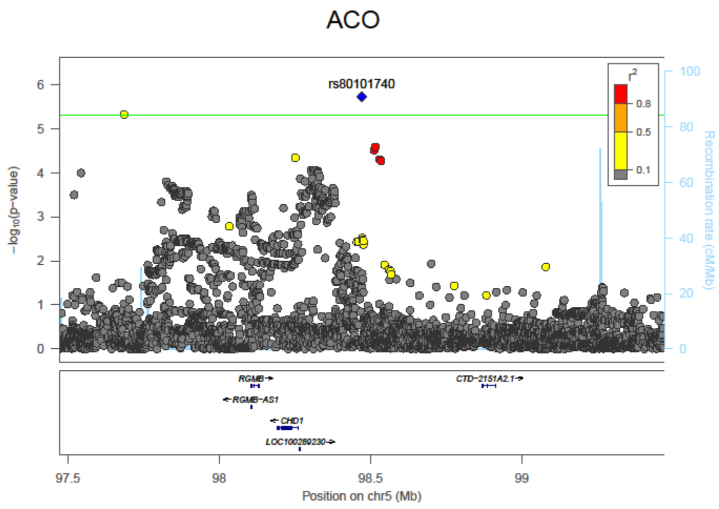


### rs35570272

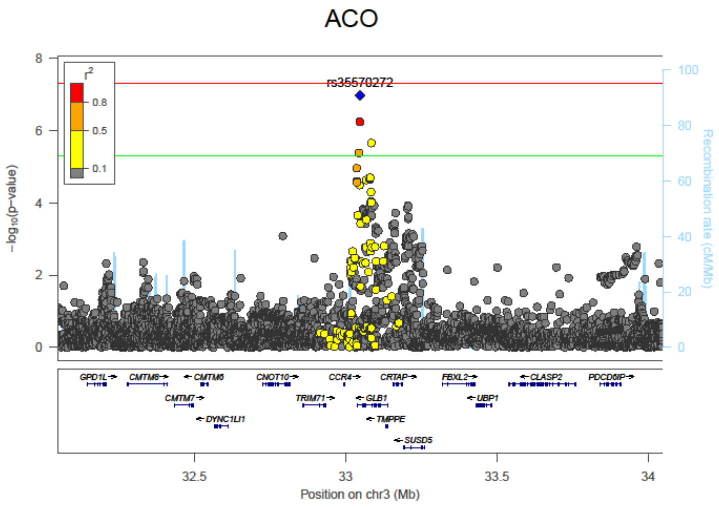

## rs16903574

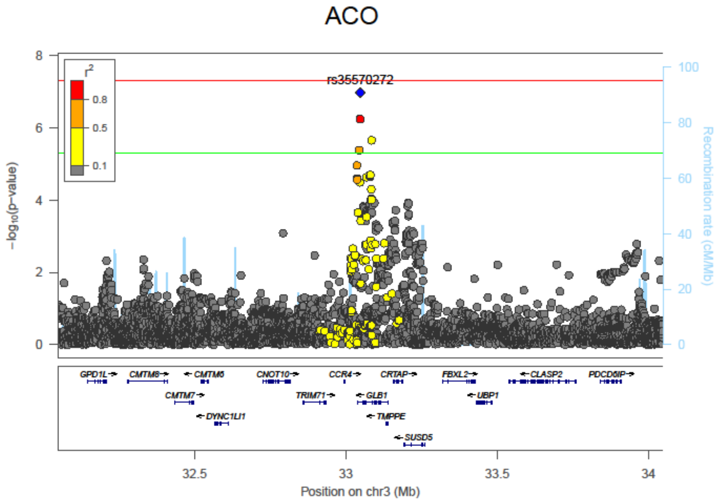


## rs2584662

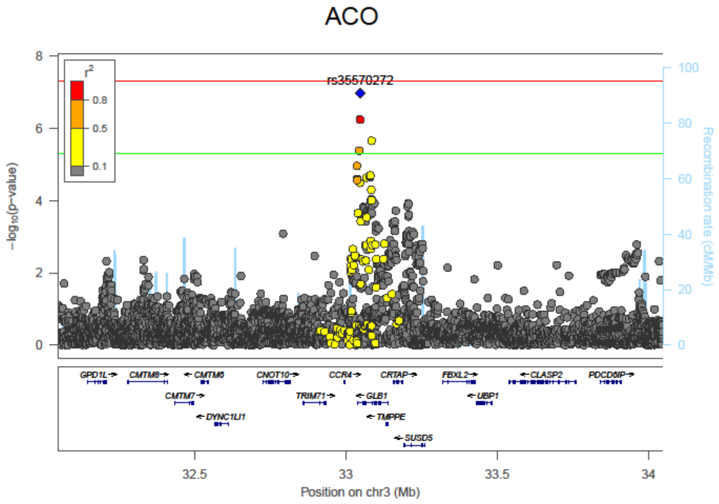


## rs1837253

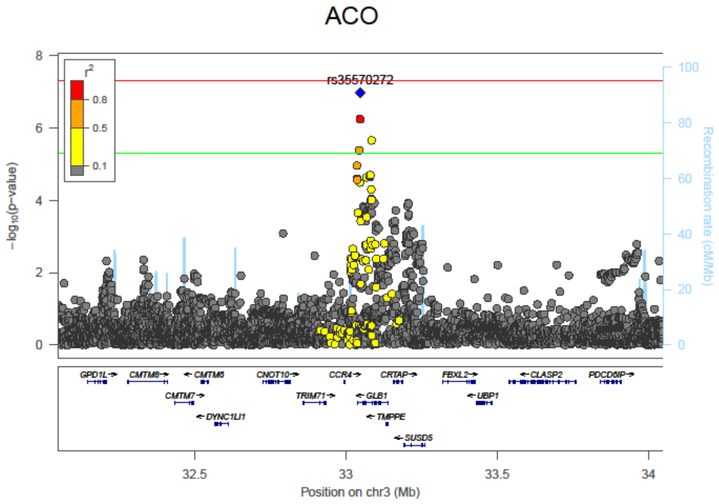

## rs6787279

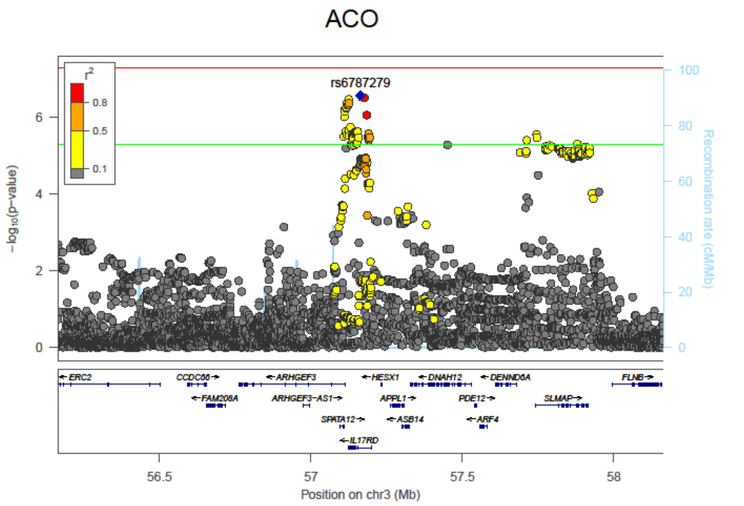


## rs9273410

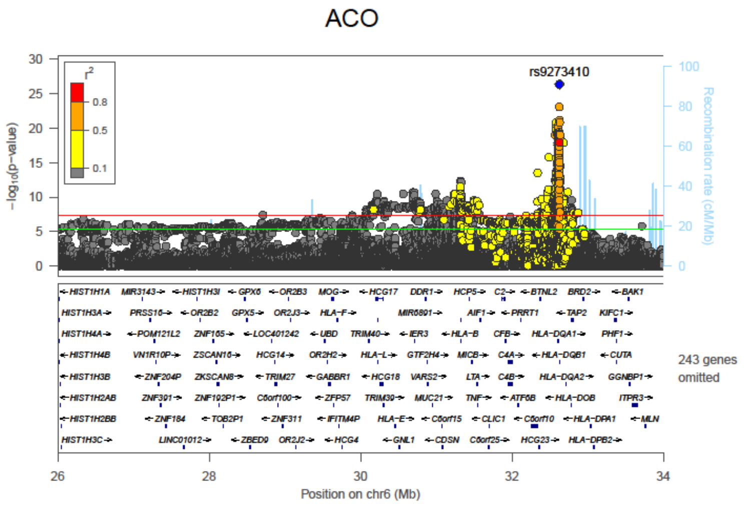


## rs3749833

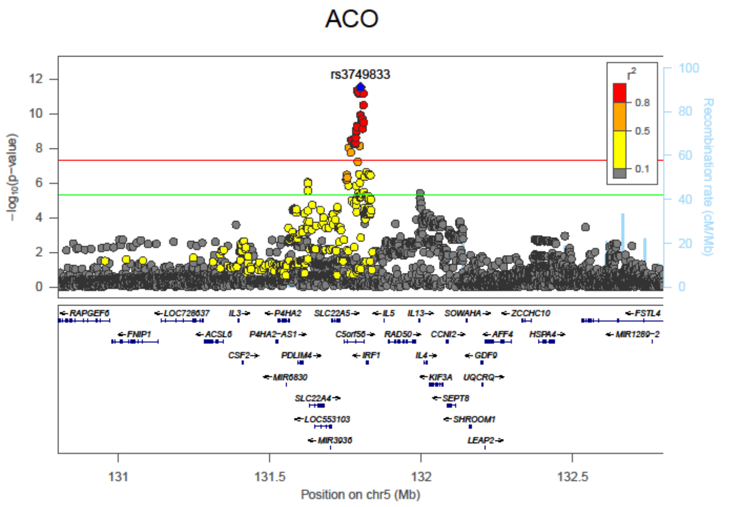

#### Study acknowledgements and funding

**Cardiovascular Health Study**: This CHS research was supported by NHLBI contracts HHSN268201200036C, HHSN268200800007C, HHSN268201800001C, N01HC55222, N01HC85079, N01HC85080, N01HC85081, N01HC85082, N01HC85083, N01HC85086; and NHLBI grants U01HL080295, R01HL087652, R01HL105756, R01HL103612, R01HL120393, and U01HL130114 with additional contribution from the National Institute of Neurological Disorders and Stroke (NINDS). Additional support was provided through R01AG023629 from the National Institute on Aging (NIA). A full list of principal CHS investigators and institutions can be found at CHS-NHLBI.org. The provision of genotyping data was supported in part by the National Center for Advancing Translational Sciences, CTSI grant UL1TR001881, and the National Institute of Diabetes and Digestive and Kidney Disease Diabetes Research Center (DRC) grant DK063491 to the Southern California Diabetes Endocrinology Research Center. The content is solely the responsibility of the authors and does not necessarily represent the official views of the National Institutes of Health.

**COPDGene:** *Grant Support and Disclaimer:* The project described was supported by Award Number U01 HL089897 and Award Number U01 HL089856 from the National Heart, Lung, and Blood Institute. The content is solely the responsibility of the authors and does not necessarily represent the official views of the National Heart, Lung, and Blood Institute or the National Institutes of Health. *COPD Foundation Funding:* The COPDGene® project is also supported by the COPD Foundation through contributions made to an Industry Advisory Board comprised of AstraZeneca, Boehringer Ingelheim, GlaxoSmithKline, Novartis, Pfizer, Siemens and Sunovion. **COPDGene® Investigators – Core Units.** *Administrative Center*: James D. Crapo, MD (PI); Edwin K. Silverman, MD, PhD (PI); Barry J. Make, MD; Elizabeth A. Regan, MD, PhD. *Genetic Analysis Center*: Terri Beaty, PhD; Ferdouse Begum, PhD; Peter J. Castaldi, MD, MSc; Michael Cho, MD; Dawn L. DeMeo, MD, MPH; Adel R. Boueiz, MD; Marilyn G. Foreman, MD, MS; Eitan Halper-Stromberg; Lystra P. Hayden, MD, MMSc; Craig P. Hersh, MD, MPH; Jacqueline Hetmanski, MS, MPH; Brian D. Hobbs, MD; John E. Hokanson, MPH, PhD; Nan Laird, PhD; Christoph Lange, PhD; Sharon M. Lutz, PhD; Merry-Lynn McDonald, PhD; Margaret M. Parker, PhD; Dmitry Prokopenko, Ph.D; Dandi Qiao, PhD; Elizabeth A. Regan, MD, PhD; Phuwanat Sakornsakolpat, MD; Edwin K. Silverman, MD, PhD; Emily S. Wan, MD; Sungho Won, PhD. *Imaging Center*: Juan Pablo Centeno; Jean-Paul Charbonnier, PhD; Harvey O. Coxson, PhD; Craig J. Galban, PhD; MeiLan K. Han, MD, MS; Eric A. Hoffman, Stephen Humphries, PhD; Francine L. Jacobson, MD, MPH; Philip F. Judy, PhD; Ella A. Kazerooni, MD; Alex Kluiber; David A. Lynch, MB; Pietro Nardelli, PhD; John D. Newell, Jr., MD; Aleena Notary; Andrea Oh, MD; Elizabeth A. Regan, MD, PhD; James C. Ross, PhD; Raul San Jose Estepar, PhD; Joyce Schroeder, MD; Jered Sieren; Berend C. Stoel, PhD; Juerg Tschirren, PhD; Edwin Van Beek, MD, PhD; Bram van Ginneken, PhD; Eva van Rikxoort, PhD; Gonzalo Vegas Sanchez-Ferrero, PhD; Lucas Veitel; George R. Washko, MD; Carla G. Wilson, MS; *PFT QA Center, Salt Lake City, UT*: Robert Jensen, PhD. *Data Coordinating Center and Biostatistics*, *National Jewish Health, Denver, CO*: Douglas Everett, PhD; Jim Crooks, PhD; Katherine Pratte, PhD; Matt Strand, PhD; Carla G. Wilson, MS. *Epidemiology Core*, *University of Colorado Anschutz Medical Campus, Aurora, CO*: John E. Hokanson, MPH, PhD; Gregory Kinney, MPH, PhD; Sharon M. Lutz, PhD; Kendra A. Young, PhD. *Mortality Adjudication Core:* Surya P. Bhatt, MD; Jessica Bon, MD; Alejandro A. Diaz, MD, MPH; MeiLan K. Han, MD, MS; Barry Make, MD; Susan Murray, ScD; Elizabeth Regan, MD; Xavier Soler, MD; Carla G. Wilson, MS. *Biomarker Core*: Russell P. Bowler, MD, PhD; Katerina Kechris, PhD; Farnoush Banaei-Kashani, Ph.D. **COPDGene® Investigators – Clinical Centers.** *Ann Arbor VA:* Jeffrey L. Curtis, MD; Perry G. Pernicano, MD. *Baylor College of Medicine, Houston, TX*: Nicola Hanania, MD, MS; Mustafa Atik, MD; Aladin Boriek, PhD; Kalpatha Guntupalli, MD; Elizabeth Guy, MD; Amit Parulekar, MD. *Brigham and Women's Hospital, Boston, MA*: Dawn L. DeMeo, MD, MPH; Alejandro A. Diaz, MD, MPH; Lystra P. Hayden, MD; Brian D. Hobbs, MD; Craig

Hersh, MD, MPH; Francine L. Jacobson, MD, MPH; George Washko, MD. *Columbia University, New York, NY*: R. Graham Barr, MD, DrPH; John Austin, MD; Belinda D'Souza, MD; Byron Thomashow, MD. *Duke University Medical Center, Durham, NC*: Neil MacIntyre, Jr., MD; H. Page McAdams, MD; Lacey Washington, MD. *Grady Memorial Hospital, Atlanta, GA: Eric Flenaugh, MD; Silanth Terpenning, MD. HealthPartners Research Institute, Minneapolis, MN*: Charlene McEvoy, MD, MPH; Joseph Tashjian, MD. *Johns Hopkins University, Baltimore, MD*: Robert Wise, MD; Robert Brown, MD; Nadia N. Hansel, MD, MPH; Karen Horton, MD; Allison Lambert, MD, MHS; Nirupama Putcha, MD, MHS. *Lundquist Institute for Biomedical Innovationat Harbor UCLA Medical Center, Torrance, CA*: Richard Casaburi, PhD, MD; Alessandra Adami, PhD; Matthew Budoff, MD; Hans Fischer, MD; Janos Porszasz, MD, PhD; Harry Rossiter, PhD; William Stringer, MD. *Michael E. DeBakey VAMC, Houston, TX*: Amir Sharafkhaneh, MD, PhD; Charlie Lan, DO. *Minneapolis VA:* Christine Wendt, MD; Brian Bell, MD; Ken M. Kunisaki, MD, MS. *National Jewish Health, Denver, CO*: Russell Bowler, MD, PhD; David A. Lynch, MB. *Reliant Medical Group, Worcester, MA*: Richard Rosiello, MD; David Pace, MD. *Temple University, Philadelphia, PA:* Gerard Criner, MD; David Ciccolella, MD; Francis Cordova, MD; Chandra Dass, MD; Gilbert D'Alonzo, DO; Parag Desai, MD; Michael Jacobs, PharmD; Steven Kelsen, MD, PhD; Victor Kim, MD; A. James Mamary, MD; Nathaniel Marchetti, DO; Aditi Satti, MD; Kartik Shenoy, MD; Robert M. Steiner, MD; Alex Swift, MD; Irene Swift, MD; Maria Elena Vega-Sanchez, MD*. University of Alabama, Birmingham, AL:* Mark Dransfield, MD; William Bailey, MD; Surya P. Bhatt, MD; Anand Iyer, MD; Hrudaya Nath, MD; J. Michael Wells, MD. *University of California, San Diego, CA*: Douglas Conrad, MD; Xavier Soler, MD, PhD; Andrew Yen, MD. *University of Iowa, Iowa City, IA*: Alejandro P. Comellas, MD; Karin F. Hoth, PhD; John Newell, Jr., MD; Brad Thompson, MD. *University of Michigan, Ann Arbor, MI:* MeiLan K. Han, MD MS; Ella Kazerooni, MD MS; Wassim Labaki, MD MS; Craig Galban, PhD; Dharshan Vummidi, MD*. University of Minnesota, Minneapolis, MN*: Joanne Billings, MD; Abbie Begnaud, MD; Tadashi Allen, MD. *University of Pittsburgh, Pittsburgh, PA*: Frank Sciurba, MD; Jessica Bon, MD; Divay Chandra, MD, MSc; Carl Fuhrman, MD; Joel Weissfeld, MD, MPH. *University of Texas Health, San Antonio, San Antonio, TX*: Antonio Anzueto, MD; Sandra Adams, MD; Diego Maselli-Caceres, MD; Mario E. Ruiz, MD; Harjinder Singh.

**ECLIPSE:** The ECLIPSE study (NCT00292552; GSK code SCO104960) was funded by GSK.

**EPIC-Norfolk:** The EPIC-Norfolk study team are grateful to all the participants who have been part of the project and to the many members of the study teams at the University of Cambridge who have enabled this research. The study is funded by Medical Research Council (MR/N003284/1, MC-UU_12015/1, MC_PC_13048) Cancer Research UK (C864/A14136).

**FHS:** This work was partially supported by the National Heart, Lung and Blood Institute's **Framingham Heart Study** (contract number N01-HC-25195) and its contract with Affymetrix, Inc for genotyping services (contract number N02-HL-6-4278). Also supported by NIH P01 AI050516.

**GASP** (moderate-severe asthma correlations): cohort collection and genotyping was funded by; Asthma UK Grant to I.S. (AUK-PG-2013-188), Rosetrees Tust (Grant to I.S.), AirPROM, U-BIOPRED (EU-IMI 115010) and an MRC Strategic Award to I.P.H., M.D.T., L.V.W. and Professor David Strachan (MC_PC_12010). Recruitment of asthma patients in Manchester was funded by the NIHR and the North West Lung Centre Charity.

**Generation Scotland** received core support from the Chief Scientist Office of the Scottish Government Health Directorates [CZD/16/6] and the Scottish Funding Council [HR03006]. Genotyping was funded by the Medical Research Council UK and the Wellcome Trust (Wellcome Trust Strategic Award "STratifying Resilience and Depression Longitudinally" (STRADL) Reference 104036/Z/14/Z). .CH is supported by an MRC University Unit Programme Grant MC_UU_00007/10

(QTL in Health and Disease). We thank all the families who took part, the general practitioners and the Scottish School of Primary Care for their help in recruiting them, and the whole Generation Scotland team, which includes interviewers, computer and laboratory technicians, clerical workers, research scientists, volunteers, managers, receptionists, healthcare assistants and nurses. Genotyping of the GS:SFHS samples was carried out by the Genetics Core Laboratory at the Edinburgh Clinical Research Facility, University of Edinburgh, Scotland.

**GenKOLS:** The Norway GenKOLS study (Genetics of Chronic Obstructive Lung Disease, GSK code RES11080) was funded by GSK.

**HUNT**: The Nord-Trøndelag Health Study (The HUNT Study) is a collaboration between HUNT Research Center (Faculty of Medicine and Health Sciences, NTNU, Norwegian University of Science and Technology), Nord-Trøndelag County Council, Central Norway Regional Health Authority, and the Norwegian Institute of Public Health. The K.G. Jebsen Center for Genetic Epidemiology is funded by Stiftelsen Kristian Gerhard Jebsen; Faculty of Medicine and Health Sciences, NTNU; The Liaison Committee for education, research and innovation in Central Norway; and the Joint Research Committee between St. Olavs Hospital and the Faculty of Medicine and Health Sciences, NTNU. The genotyping in HUNT was financed by the National Institute of Health (NIH); University of Michigan; The Research Council of Norway; The Liaison Committee for education, research and innovation in Central Norway; and the Joint Research Committee between St. Olavs Hospital and the Faculty of Medicine and Health Sciences, NTNU.

**Lovelace Smokers Cohort**: The cohort and analysis was primarily supported by National Cancer Institute grant R01 CA097356 (SAB). The State of New Mexico as a direct appropriation from the Tobacco Settlement Fund to SAB through collaboration with University of New Mexico provided initial support to establish the LSC. Additional support was provided by NIH/NCI P30 CA118100 (SAB), HL68111 (Y.T.), and HL107873-01 (YT and SB).

**Rotterdam Study:** The study is funded by Erasmus Medical Center and Erasmus University, Rotterdam, Netherlands Organization for the Health Research and Development (ZonMw), the Research Institute for Diseases in the Elderly (RIDE), the Ministry of Education, Culture and Science, the Ministry for Health, Welfare and Sports, the European Commission (DG XII), and the Municipality of Rotterdam. The generation and management of GWAS genotype data for the Rotterdam Study (RS I, RS II, RS III) was executed by the Human Genotyping Facility of the Genetic Laboratory of the Department of Internal Medicine, Erasmus MC, Rotterdam, The Netherlands. The GWAS datasets are supported by the Netherlands Organisation of Scientific Research NWO Investments (nr. 175.010.2005.011, 911-03-012), the Genetic Laboratory of the Department of Internal Medicine, Erasmus MC, the Research Institute for Diseases in the Elderly (014-93-015; RIDE2), the Netherlands Genomics Initiative (NGI)/Netherlands Organisation for Scientific Research (NWO) Netherlands Consortium for Healthy Aging (NCHA), project nr. 050-060-810. The generation and management of spirometric data was supported by FWO project G035014N.

**SPIROMICS:** The Subpopulations and Intermediate Outcomes in COPD Study was supported by contracts from the NIH/NHLBI (HHSN268200900013C, HHSN268200900014C, HHSN268200900015C, HHSN268200900016C, HHSN268200900017C, HHSN268200900018C, HHSN268200900019C, HHSN268200900020C), which were supplemented by contributions made through the Foundation for the NIH from AstraZeneca; Bellerophon Therapeutics; Boehringer-Ingelheim Pharmaceuticals, Inc; Chiesi Farmaceutici SpA; Forest Research Institute, Inc; GSK; Grifols Therapeutics, Inc; Ikaria, Inc; Nycomed GmbH; Takeda Pharmaceutical Company; Novartis Pharmaceuticals Corporation;

Regeneron Pharmaceuticals, Inc; and Sanofi. We acknowledge all investigators, staff, and participants in SPIROMICS.
